# Spanish-version of the Posttraumatic Stress Disorder Checklist for DSM-5 (PCL-5) in health-workers

**DOI:** 10.1101/2025.05.29.25328607

**Authors:** David Villarreal-Zegarra, Kelly De la Cruz-Torralva, Juan Barrera-Begazo, Jaime Rosales-Rimache, Percy Soto-Becerra, Ana L. Vilela-Estrada, Cristian Diaz, Moises Apolaya, Rafael Lea, C. Mahony Reategui-Rivera, Felipe E. García

## Abstract

**Background:** The Posttraumatic Stress Disorder Checklist (PCL-5) is one of the most widely used instruments for assessing posttraumatic stress disorder (PTSD), adapted to meet DSM-5 criteria. Establishing a validated online version for Spanish-speaking populations, particularly among healthcare professionals, is essential to ensure accurate assessment aligned with these updated criteria.

**Aim:** The purpose is to evaluate the psychometric properties of the online Spanish version of the PCL-5 for healthcare professionals exposed to the COVID-19 health emergency.

**Methods:** The study involved 1543 Peruvian healthcare professionals aged 20 to 70 recruited through a virtual platform. In addition to the PCL-5, instruments were administered to assess depressive symptoms (assessed by PHQ-9) and anxious symptoms (assessed by GAD-7).

**Results:** The results showed that the 7-factor model presented the best fit among all models (CFI=0.967; TLI=0.958; SRMR=0.030; RMSEA[CI90%]=0.062 [0.057-0.067]) and met the measurement invariance across sexes, age groups, and educational levels, with adequate values of internal consistency (ΔCFI<0.01; ΔRMSEA<0.01). Moreover, a high latent correlation was observed with depressive and anxious symptoms (≥0.60).

**Conclusion:** The online Spanish version of the PCL-5 scale is a valid and reliable tool for assessing PTSD among Spanish-speaking healthcare professionals, representing the only instrument adapted to DSM-5 criteria for this population.

## Background

PTSD diagnosis traditionally follows the criteria outlined in the Diagnostic and Statistical Manual of Mental Disorders (DSM). However, these criteria have been modified in the last decade, reflected in the changes observed between DSM-IV and DSM-5. In the updated version, individual response to the event (intense fear, helplessness, or horror) was removed, and special emphasis was placed on the behavioral symptoms that accompany PTSD. The current model proposes four diagnostic criteria defined as re-experiencing (recurrent and distressing thoughts about the traumatic event), avoidance (avoidance of situations or thoughts related to the trauma), negative cognitions (negative thoughts, guilt, fear, and difficulty experiencing positive emotions) and mood, and arousal (irritability, difficulty concentrating, and exaggerated responses to stimuli), as well as emphasizing the "fight" and "flight" aspects associated with PTSD. Finally, the distress of the disorder must be experienced continuously for more than one month (American Psychiatric Association, 2014).

Various diagnostic tools, such as the Posttraumatic Diagnostic Scale for DSM-5 (PDS-5) (Foa et al., 2016), and the Dissociative Subtype of PTSD Scale (DSPS) (Wolf et al., 2017), aid in PTSD diagnosis. In this study, we focus on the PTSD Checklist for DSM-5 (PCL-5), an updated DSM-5 instrument widely used for the assessment of adults at risk for PTSD; the choice of the PCL-5 is justified by its ease of use, its adaptability to different contexts of and its ability to provide a rapid and reliable assessment of PTSD symptoms (Armour et al., 2016). Assessing the degree to which the presence and severity of symptoms bother you (Brewin, 2005). The latest version adapts from a three-factor model with 17 symptoms to a four-factor model with 20 symptoms.

There are several heterogeneous models of the PCL-5; the original model based on the DSM-5 (four dimensions/ re-experiencing, avoidance, negative cognitions and mood, and arousal) gives rise to the Externalizing Behavior Model (six dimensions/avoidance behavior, irritability, impulsivity, substance use, sensation seeking behavior and aggression) (Tsai et al., 2015), the Anhedonia Model (six dimensions/ intrusion, avoidance, negative affect, anhedonia, dysphoric arousal, and anxious arousal) (Liu et al., 2014), and the Seven Factor Model (seven dimensions/ re-experiencing, avoidance, negative affect, anhedonia, externalizing behaviors, and anxious and dysphoric arousal) (Armour et al., 2015). From DSM-IV, the DSM-IV Model (three dimensions/intrusion, avoidance, and hyperarousal) (Bliese et al., 2008), the DSM-IV Dysphoria Model (four dimensions/intrusion, avoidance, dysphoria and hyperarousal) (Simms et al., 2002), and the DSM-IV Dysphoria Arousal Model (five dimensions/ intrusion, avoidance, hyperarousal, dysphoria and Dysphoric Arousal) (Elhai et al., 2011). These various models reflect the complex and multifaceted factorial structure of PTSD, which has required diverse modeling approaches depending on sample characteristics, cultural context, and methodology used (Armour et al., 2016; Ashbaugh et al., 2016; Caldas et al., 2020; Schmitt et al., 2018).

The PCL-5 has been extensively studied in clinical populations; however, research among healthcare workers remains limited (Cheng et al., 2020; Morrison et al., 2021). Healthcare workers are frequently exposed to high levels of stress and traumatic events in the workplace, as exemplified by the COVID-19 pandemic. The emergence of SARS-CoV-2 and the subsequent COVID-19 pandemic significantly affected health, well-being, and quality of life (Ruiz et al., 2020). Frontline healthcare workers were among the most impacted occupational groups, facing heightened infection risk and subsequent mental distress, leading to mood disorders and posttraumatic stress disorder (PTSD) (Krishnamoorthy et al., 2020; Lai et al., 2020; Luo et al., 2020; Xiong et al., 2020). During this health crisis, one in four healthcare professionals worldwide suffered from depression and/or anxiety problems (Sahebi et al., 2021) due to increased workload, shortage of personal protective equipment, physical isolation, stigma, personal losses, limited resources, stress, and concern about being infected (Dosil Santamaría et al., 2021; Mokhtari et al., 2020; Pfefferbaum & North, 2020; Rabow et al., 2021; Shaukat et al., 2020; Singh & Subedi, 2020; Zhou et al., 2020), among other causes. Therefore, the pandemic exacerbated the risk of developing PTSD among healthcare workers (Carmassi et al., 2020).

Although the online version of the PCL-5 has been validated for multiple populations, including undergraduate students and frontline healthcare workers (Ashbaugh et al., 2016; Cheng et al., 2020), a validated online Spanish version of the PCL-5 specifically tailored to healthcare workers is still lacking. With the changes in the DSM-5 and the critical need to assess PTSD in healthcare workers exposed to the COVID-19 pandemic, a reliable and valid assessment instrument is essential. This study, therefore, seeks to evaluate the psychometric properties of the PCL-5 among Spanish speaker’s healthcare professionals who were on the front lines during the COVID-19 health crisis.

## Methods

### Desing

Our study was cross-sectional, and we used the STROBE Checklist (see supplementary material 1). This study is part of a primary study “FRONTLINE Study”, led by Centro Nacional de Salud Ocupacional y Protección del Ambiente para la Salud of Instituto Nacional de Salud (INS) and Instituto de Evaluación de Tecnologías de la Salud e Investigación (IETSI) at EsSalud.

### Participants and Setting

The study focused on healthcare professionals who worked in health facilities in Peru during the COVID-19 pandemic. We included the five most common occupational groups in the Peruvian healthcare system: physicians, nurses, midwives, medical technicians, and health technicians. The evaluation encompassed workers in facilities affiliated with the Ministry of Health, regional governments, or social health insurance (EsSalud). We excluded participants with missing data on the PCL-5 and psychometric scales used.

During the pandemic, Peru was one of the countries with the highest number of COVID-19 deaths worldwide (Johns Hopkins University, 2022). In addition, the Peruvian health system is fragmented, both in terms of coverage and service delivery. The Peruvian healthcare workers are spread across four health providers: the Ministry of Health, EsSalud, the Armed Forces and Police, and the private health system. As a result, working conditions and workloads vary (Carrillo-Larco et al., 2022).

The sampling was non-probabilistic. Recruitment was conducted through direct invitations sent to healthcare professionals listed with EsSalud and INS via email, WhatsApp, and institutions related to healthcare work. Additionally, dissemination was carried out through social networks, including Twitter and Facebook, between December 21, 2021, and August 31, 2022. The REDCap program was used to design and collect data from the participants who responded to the anonymous, online, voluntary survey. Participants had to validate a captcha code to give informed consent. The data were stored on IETSI-EsSalud’s local servers, which have the institutional license for REDCap.

The minimum number of participants was determined to be 463, based on a Comparative Fit Index (CFI) of 0.95, seven dimensions (with 5, 2, 4, 3, 2, 2, and 2 items, respectively), an average factor loading of 0.5, a latent correlation between dimensions of 0.7, a two-tailed alpha level of 0.05, a power of 80%, and assuming 149 degrees of freedom.

### Instruments

#### The online version of PTSD Checklist for DSM-5 (PCL-5)

The PCL-5 is a self-report instrument that assesses 20 symptoms of posttraumatic stress disorder (PTSD) according to DSM-5 criteria (Blevins et al., 2015). Respondents rate the degree to which each symptom affects them on a five-point Likert scale (0 = not at all; 4 = extremely). The PCL-5 assesses the four indicators of PTSD proposed in the DSM-5 through the following items: Intrusion (questions 1-5), Avoidance (questions 6-7), Negative Disturbances in Cognition and Mood (questions 8-14), and Disturbances in Symptomatic Arousal and Reactivity (questions 15-20).

One of the ways to meet the DSM-5 criteria for PTSD, respondents’ PCL-5 scores must follow the following rules (Blevins et al., 2015): (a) A total score at or above the cut-off (≥33 points); (b) Each item rated as 2 = "Moderately" or higher as an endorsed symptom, then meet the minimum number of symptomatic items required by the DSM-5 diagnostic rule in the different dimensions (i.e., at least one item from the Intrusion dimension, one item from Avoidance, two items from Negative Disturbances in Cognition and Mood, and two items from Disturbances in Symptomatic Arousal and Reactivity).

The internal consistency of the PCL-5 total score has been reported to be 0.90-0.96 (for the four subscales: intrusions: α = 0.77 - 0.92; avoidance: α = 0.74 - 0.92; negative cognitive and mood disturbances: α = 0.78 - 0.89; hyperarousal: α = 0.75 - 0.84), also, test-retest reliability reported values between 0.66 and 0.91 (Blevins et al., 2015).

#### Patient Health Questionnaire (PHQ-9)

The PHQ-9 is a nine-item self-report Likert-type questionnaire(Spitzer et al., 1999). These items were developed based on the nine criteria used in the DSM-IV to assess major depressive disorder (MDD), criteria that are retained in the DSM-5. The PHQ-9 has four response options: 0 ‘never,’ 1 ‘several days,’ 2 ‘more than half the days’, and 3 ‘almost every day,’ resulting in scores ranging from 0 to 27. It aims to measure indicators of depressive symptoms experienced in the past two weeks.

The PHQ-9 shows adequate evidence of one-dimension validity in the Peruvian context (CFI = 0.94; TLI = 0.91; RMSEA = 0.09; SRMR = 0.04) (Villarreal-Zegarra et al., 2019). In addition, optimal reliability values were found through internal consistency coefficients (α = 0.87; ω = 0.87) (Villarreal-Zegarra et al., 2019). The PHQ-9 has a specificity of 54.2 (48.7 - 59.6) and a specificity of 87.4 (85.2 - 89.3) in the Peruvian population for a cut-off of 10 points or more being as the gold standard a diagnosis of major depression based on a psychiatric interview (Villarreal-Zegarra et al., 2023).

#### Generalized Anxiety Disorder (GAD-7)

The GAD-7 is a measure of anxiety that consists of seven questions using a four-point Likert scale ranging from 0 (not at all) to 3 (almost every day) (Spitzer et al., 2006). The total score ranges from 0 to 21, with higher scores indicating higher anxiety symptoms. This instrument assesses seven anxiety symptoms based on the DSM-IV, related to the frequency and severity of distress experienced in the past two weeks (Franco-Jimenez & Nuñez-Magallanes, 2022).

The one-dimensional model of the instrument shows adequate psychometric properties in Peru (GFI = 0.96; AGFI = 0.91; CFI = 0.99; NFI = 0.98; RMSEA = 0.08) (García-Campayo et al., 2010). The GAD-7 has a specificity of 39.3 (21.5 - 59.4) and a specificity of 88.4 (86.5 - 90.1) in the Peruvian population for a cut-off of 10 points or more being as the gold standard a diagnosis of generalized anxiety based on a psychiatric interview (Villarreal-Zegarra et al., 2023).

#### Covariates

Our covariates were sex (male, female), age groups (young adults [20 to 29 years], middle adulthood [30 to 45 years], and established adults [46 to 70 years]), education levels (technical professional, university undergraduate, and university postgraduate), profession (physician, nurse, obstetrician, medical technologist, and assistance technician), work modality (face-to-face, remote, mixed, dońt know), and marital status (single, married, divorced, cohabited, other).

### Analysis plan

First, we conducted a descriptive analysis based on the socio-characteristics of the participants. Second, we performed a confirmatory factor analysis using the Maximum Likelihood Robust estimator (Suh, 2015) and Pearson’s matrices. The model was evaluated around four goodness-of-fit indices: Comparative Fit Index (CFI) and Tucker-Lewis Index (TLI), which should have values greater than 0.95, and Root Mean Square Error of Approximation (RMSEA) and Standardized Root Mean Square Residual (SRMR), which should have values less than 0.08 (Sun, 2005). Also, we use the Akaike information criterion (AIC) and Bayes information criterion (BIC), selecting the model with the lowest value. We selected the best-fitting model by identifying which had adequate CFI, TLI, RMSEA, and SRMR values, and then saw which had the lowest AIC and BIC. Third, a multigroup factor analysis (invariance) was performed based on sex, age group, and education level. We use Wu and Estabrook correction (Wu & Estabrook, 2016) and theta parameterization. We considered that there was invariance when the comparison between the different models showed ΔCFI values of less than 0.01 (Putnick & Bornstein, 2016). Fourth, we evaluated the internal consistency based on the alpha and omega coefficients, considered optimal values when they are greater than 0.80, and adequate when they are greater than 0.70 (Kalkbrenner, 2023). Fifth, convergent validity was evaluated using latent correlation coefficients derived from a structural equation modeling analysis, assessing the relationships between the PCL-5 and the PHQ-9 (measuring depressive symptoms) as well as the GAD-7 (measuring anxious symptoms). For the interpretation of these coefficients, we used the following categories: weak (r > 0.10), moderate (r > 0.30) or strong (r > 0.50) according to the size of the correlation (Akoglu, 2018).

### Ethics topics

Our research was based on the analysis of secondary data from the study titled "Peruvian Frontline HealthcaRe wOrkers meNTal heaLth Issues during paNdEmics: The Peruvian FRONTLINE Study". This study was reviewed and approved by the Institutional Ethics and Research Committee of the Peruvian National Institute of Health (Review code: OC-029-2021). The Peruvian FRONTLINE study aims to perform a nationwide assessment of the mental health of Peruvian healthcare workers during the pandemic. Participation in the study was voluntary, and written informed consent was obtained through an online questionnaire.

## Results

### Participant’s characteristics

A total of 1543 people aged 18 years and older, working in public or private health facilities in Peru between December 2021 and August 2022, participated. Of the total, 71.9% were women (n = 1110) and 28.1% men (n = 433). The average age was 46.5 years, ranging from 20 to 70 years. Most had postgraduate studies (54.5%; n=841) and single marital status (42.6%; n=657). Some 31.3% were nurses, 29.9% were physicians, 18.8% technical assistants, 13.3% obstetricians and 6.8% medical technologists. In addition, 6.4% self-reported a previous diagnosis of depression and 5.8% of anxiety. Table 1 includes complete socio-demographic characteristics. The total sample consisted of 1999 records; however, 456 were excluded because of missing data on the variables of interest.

**Figure 1.**
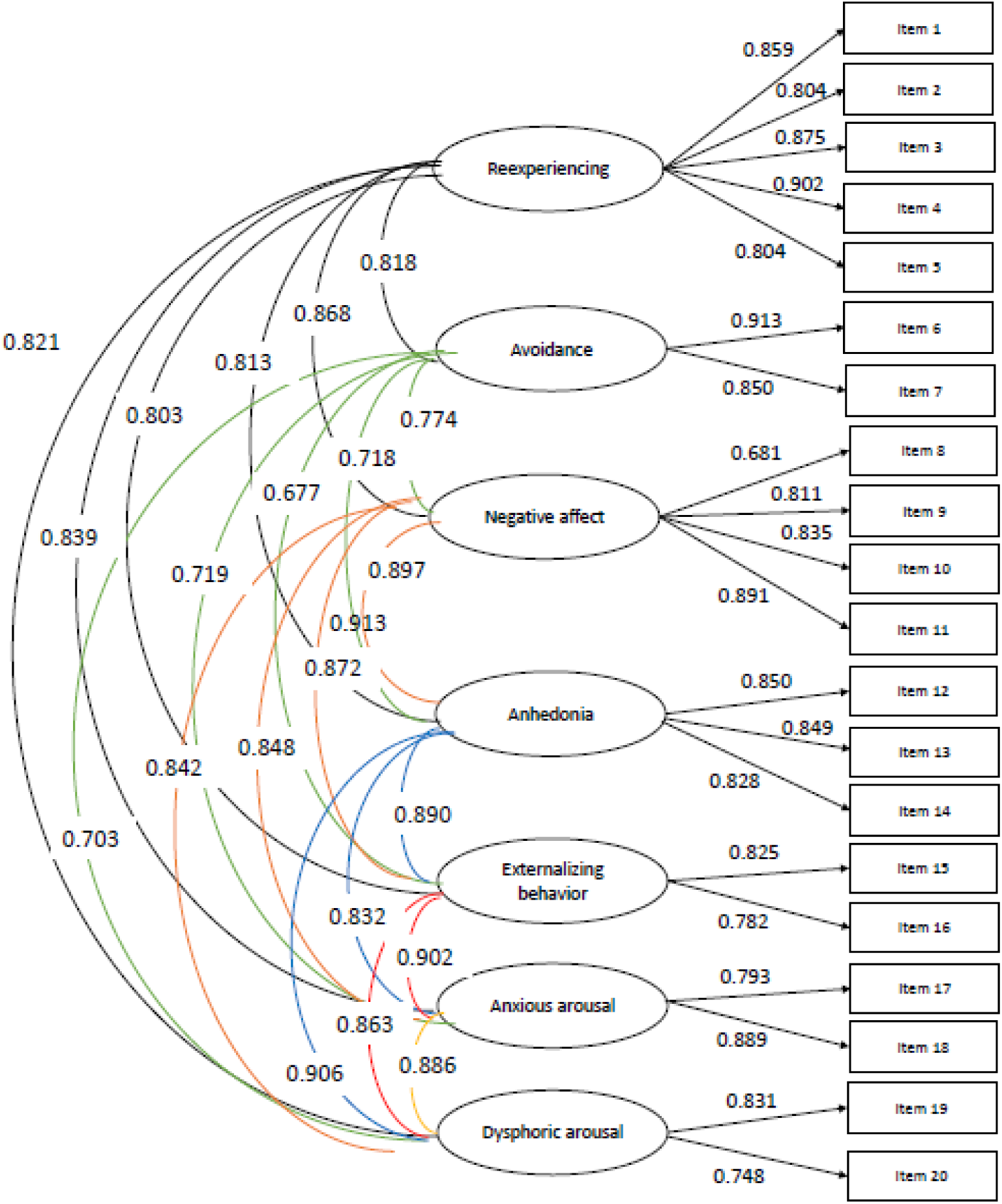
Factor structure of the PCL-5.

**Table 1.** Sociodemographic characteristics (n = 1543)

|  | n | % |
| --- | --- | --- |
| Sex |  |  |
| Female | 1110 | 71.9% |
| Male | 433 | 28.1% |
| Age |  |  |
| Young adults (20 - 29) | 245 | 15.9% |
| Middle adulthood (30 - 45) | 468 | 30.3% |
| Established adults (46 - 70) | 830 | 53.8% |
| Education level |  |  |
| Technical professional | 270 | 17.5% |
| University undergraduate | 432 | 28.0% |
| University postgraduate | 841 | 54.5% |
| Profession |  |  |
| Physician | 461 | 29.9% |
| Nurse | 483 | 31.3% |
| Obstetrician | 204 | 13.2% |
| Medical Technologist | 105 | 6.8% |
| Assistance Technician | 290 | 18.8% |
| Depression diagnosis based on PHQ-9 |  |  |
| Yes | 99 | 6.4% |
| No | 1444 | 93.6% |
| Diagnostic anxiety based on GAD-7 |  |  |
| Yes | 90 | 5.8% |
| No | 1453 | 94.2% |
| Work modality |  |  |
| Face-to-face modality | 1264 | 81.9% |
| Remote modality | 48 | 3.1% |
| Mixed modality | 30 | 1.9% |
| Don't know | 201 | 13.0% |
| Live alone |  |  |
| Yes | 240 | 15.6% |
| No | 1139 | 73.8% |
| Don't know | 164 | 10.6% |
| Marital status |  |  |
| Single | 657 | 42.6% |
| Married | 534 | 34.6% |
| Divorced | 57 | 3.7% |
| Cohabited | 248 | 16.1% |
| Other | 47 | 3.0% |
| Zone |  |  |
| No Lima | 940 | 60.9% |
| Lima | 603 | 39.1% |

Note: PHQ-9 = depressive symptoms. GAD-7 = depressive symptoms.

### Internal structure

The one-dimensional model was excluded due to goodness-of-fit indices falling significantly below recommended thresholds. Additionally, the bifactor models failed to converge and were thus not considered further. The remaining multidimensional and second-order models of the PCL-5 demonstrated acceptable goodness-of-fit indices. Consequently, AIC and BIC criteria were applied, with the seven-factor multidimensional model showing the optimal fit (CFI = 0.967; TLI = 0.958; SRMR = 0.030; RMSEA [CI 90%] = 0.062 [0.057-0.067]). Table 2 presents the goodness-of-fit indices for each evaluated model.

**Table 2.** Análisis factorial confirmatorio de los modelos del PCL-5 (n = 1556).

| Models | X<sup>2</sup> | gl | CFI | TLI | SRMR | RMSEA[CI90%] | AIC | BIC |
| --- | --- | --- | --- | --- | --- | --- | --- | --- |
| Correlated model |  |  |  |  |  |  |  |  |
| One-dimension model | 1373.2 | 90 | 0.870 | 0.848 | 0.052 | 0.137 [0.131-0.143] | 43449.4 | 43609.6 |
| Original model bases on DSM-5 (4 dimensions) | 859.4 | 164 | 0.949 | 0.941 | 0.034 | 0.073 [0.068-0.078] | 56165.7 | 56411.4 |
| Externalizing behavior model (6 dimensions) | 765.7 | 155 | 0.955 | 0.945 | 0.032 | 0.070 [0.065-0.075] | 55994.2 | 56288.0 |
| Anhedonia model (6 dimensions) | 654.5 | 155 | 0.964 | 0.955 | 0.032 | 0.063 [0.058-0.068] | 55770.2 | 56063.9 |
| Sevenfactor model (7 dimensions)* | 605.3 | 149 | 0.967 | 0.958 | 0.030 | 0.062 [0.057-0.067] | 55681.7 | 56007.5 |
| DSM-IV model (3 dimensions) | 1338.2 | 167 | 0.913 | 0.901 | 0.041 | 0.094 [0.090-0.099] | 57114.6 | 57344.2 |
| DSM-IV dysphoria model (4 dimensions) | 901.8 | 164 | 0.946 | 0.937 | 0.035 | 0.075 [0.071-0.080] | 56250.1 | 56495.9 |
| DSM-IV dysphoricarousal (5 dimensions) | 809.4 | 160 | 0.952 | 0.943 | 0.033 | 0.071 [0.067-0.076] | 56074.1 | 56341.2 |
| Second Order Model |  |  |  |  |  |  |  |  |
| Sevenfactor model with high-order (7 dimensions) | 767.4 | 163 | 0.956 | 0.948 | 0.038 | 0.068 [0.063-0.073] | 55984.5 | 56235.5 |
| Bifactor |  |  |  |  |  |  |  |  |
| Original model bases on DSM-5 with global dimension | not converge |  |  |  |  |  |  |  |
| Sevenfactor model with global dimension | not converge |  |  |  |  |  |  |  |

Note: CFI and TLI = comparative adjustment index considers optimal values $\geq 0.95$ ; SRMR = Mean root of standardized error considers adequate values $< 0.08$ ; RMSEA = Mean root of the approximation error considers appropriate values $< 0.08$ . AIC and BIC are chosen with the model with the lowest value. \* The best fitting model.

### Measurement invariance

We analyzed measurement invariance using the seven-factor multidimensional model (see Table 3). We found that measurement invariance was met between gender, age group, and educational level (ΔCFI<0.01). Therefore, comparisons can be made between these groups.

**Table 3.** Measurement invariance in the PCL-5 for the different groups.

| Group | Invariance | X<sup>2</sup> | df | CFI | TLI | SRMR | RMSEA [CI90%] | ΔCFI | ΔRMSEA |
| --- | --- | --- | --- | --- | --- | --- | --- | --- | --- |
| Sex | Configural | 802.418 | 298 | 0.965 | 0.955 | 0.032 | 0.064 [0.059-0.069] | - | - |
|  | Metric | 816.858 | 311 | 0.965 | 0.957 | 0.036 | 0.063 [0.057-0.068] | 0.000 | 0.001 |
|  | Strong | 847.436 | 324 | 0.964 | 0.958 | 0.036 | 0.062 [0.057-0.067] | 0.001 | 0.001 |
| Age group | Configural | 1001.942 | 447 | 0.964 | 0.954 | 0.034 | 0.064 [0.059-0.070] | - | - |
|  | Metric | 1038.313 | 473 | 0.963 | 0.955 | 0.041 | 0.063 [0.058-0.068] | 0.001 | 0.001 |
|  | Strong | 1082.107 | 499 | 0.963 | 0.957 | 0.042 | 0.062 [0.057-0.067] | 0.002 | 0.001 |
| Education level | Configural | 1066.905 | 447 | 0.960 | 0.949 | 0.034 | 0.068 [0.063-0.073] | - | - |
|  | Metric | 1095.748 | 473 | 0.960 | 0.951 | 0.041 | 0.067 [0.061-0.072] | 0.000 | 0.001 |
|  | Strong | 1154.265 | 499 | 0.958 | 0.952 | 0.042 | 0.066 [0.061-0.071] | 0.002 | 0.001 |

Note: CFI and TLI = comparative adjustment index considers optimal values $\geq 0.95$ ; SRMR = Mean root of standardized error considers adequate values $< 0.08$ ; RMSEA = Mean root of the approximation error considers appropriate values $< 0.08$ .

### Internal consistency

Five of the PCL-5 showed optimal values for their internal consistency coefficients: Re-experiencing (α=0.92; ω=0.92), Avoidance (α=0.90; ω=0.90), Negative affect (α=0.87; ω=0.88), Anhedonia (α=0.87; ω=0.88), and Anxious arousal (α=0.82; ω=0.82). In addition, the dimensions of Externalising Behaviour (α=0.77; ω=0.78) and Dysphoric Arousal (α=0.76; ω=0.76) showed adequate internal consistency values (see Table 4).

**Table 4.**
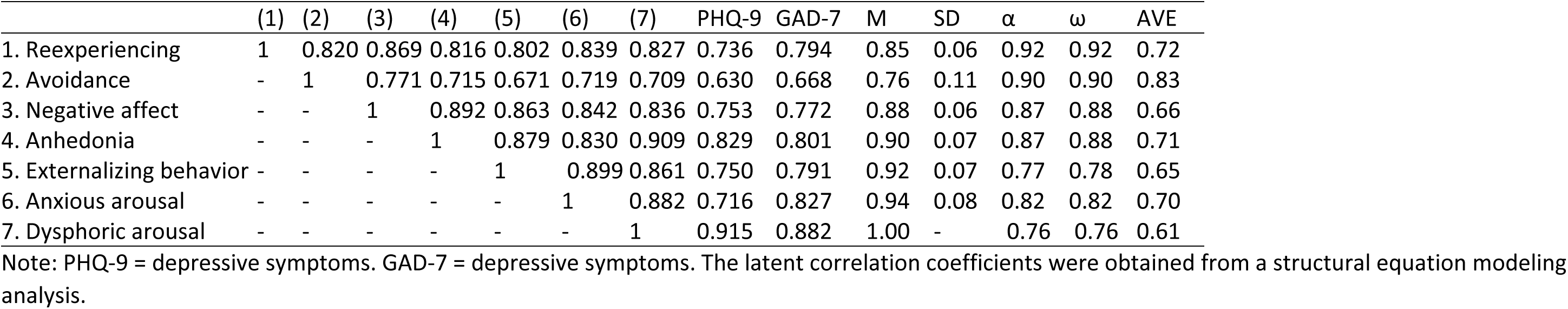
Latent correlation of PCL-5 with PHQ-9, GAD-7, and internal consistency analysis.

### Relationship with other variables

Our study found a high correlation (>0.60) between the seven dimensions of the PCL-5 and depressive and anxious symptoms. This suggests that the dimensions are closely related (see Table 4).

## Discussion

### Main findings and interpretations

Our study successfully provided evidence of validity and reliability for the online version of the Posttraumatic Stress Disorder Checklist for DSM-5 (PCL-5) among Spanish-speaking healthcare workers, supporting its application in this context. The findings identified that the Hybrid Model of seven factors best fits all models despite the four-factor model being based on the DSM-5. The adaptations made in various populations, including university students (Durón-Figueroa et al., 2019), military personnel (Bovin et al., 2016), healthcare workers (Cheng et al., 2020), traumatized populations (Ito et al., 2019), and community samples (Seligowski & Orcutt, 2016), support that the Hybrid Model has a superior fit (Armour et al., 2015). Thus, this study expanded the evidence of the seven-factor model to a sample of healthcare workers.

In addition, the instrument demonstrated the presence of configurational, metric, and scalar measurement invariance between sexes, age groups, and educational levels. This means that this instrument can be used as a screening tool for healthcare personnel, allowing for an equivalent evaluation of a heterogeneous group of health professionals between the different sexes, as indicated by a study in the Italian population (Di Tella et al., 2022). Regarding internal consistency, the instrument showed an optimal value for all subscales of the PCL-5. In contrast, the subscales "externalizing behavior" and "arousal dysphoria" reported an adequate internal consistency value for the Peruvian population.

Our study found a high correlation between the seven dimensions of PCL-5 and depressive symptoms (PHQ-9) and anxious symptoms (GAD-7). This suggests that the dimensions are closely related. Several studies have documented the high comorbidity between PTSD, anxiety, and depression, showing that patients with PTSD have a higher chance of developing depressive and anxious symptoms and that these symptoms may be a risk factor for the persistence of PTSD (Grisanzio et al., 2018; Marshall, 2020; Wu et al., 2022). Therefore, the findings of our study support the importance of addressing these disorders holistically and considering the presence of PTSD symptoms in patients with depression and anxiety.

This research shows that the model with the best fit is the 7-factor hybrid model, which combines characteristics of the Externalizing behavior model (Tsai et al., 2015) and Anhedonia model (Liu et al., 2014), both of the six factors. The hybrid model includes the following dimensions: re-experience, avoidance, negative affect, anhedonia, externalized behavior, anxious arousal, and dysphoric activation, which is based on theoretical evidence proposing that hyperarousal should consist of anxious and dysphoric arousal, that negative and positive affect are two distinct constructs; and that externalizing behaviors characterized by difficulties with emotion regulation and impulse control would be distinct from other PTSD symptoms representing passive experiences, thoughts, and feelings (Armour et al., 2015; Friedman, 2013).

An inherent limitation of the 7-factor model is that four factors have only two indicators, leading to estimation instability and other statistical problems (Armour et al., 2016). However, it is essential to note that several of the other suggested models of PTSD, including the original four-factor DSM-5 model, are composed of two items (for example, the avoidance factor). To overcome these possible limitations, a sample of at least 400 participants should be considered, which may allow for a completely valid and replicable solution (Armour et al., 2015). This research fulfills this suggestion since it works with a sample of 1543 people.

### Public health implications

Researchers and mental health professionals worldwide use the PCL-5 to assess PTSD symptoms according to DSM-5 criteria (Armour et al., 2016). Therefore, this study offers the possibility of having a valid and reliable tool to diagnose PTSD in the Peruvian population, which is of utmost importance for the public health of the country, as it would enable early identification and timely treatment.

It is essential for Peru, given its history of political and social violence, where many individuals may have experienced significant trauma. The validation of the PCL-5 for the Peruvian context would allow for a better understanding of the prevalence of PTSD and its impact on the mental health of the population, information that could be used for the development of more effective programs and public policies for the treatment and prevention of the disorder.

### Strengths and Limitations

The primary strength of this study lies in being the first to evaluate the online version of the PCL-5, specifically among Spanish-speaking healthcare workers. However, the lack of other DSM-5-adapted scales limited the ability to establish the questionnaire’s concurrent validity. Also, the absence of a gold standard precluded the assessment of the questionnaire’s sensitivity and specificity and the determination of an appropriate cut-off score. Additionally, the use of non-probabilistic sampling restricts the generalizability of the findings to the broader population of Spanish-speaking healthcare workers. Future research should focus on establishing measurement invariance across professional groups and evaluating the instrument’s sensitivity and specificity. Complementing these efforts with clinical interviews to confirm diagnoses based on DSM-5 criteria would further enhance the validity of the findings.

### Conclusions

This study demonstrates that the seven-factor model exhibited the best fit among all tested models and achieved measurement invariance across gender, age groups, and educational levels while displaying adequate internal consistency. Furthermore, the scale showed a strong correlation with depressive symptoms (PHQ-9) and anxiety symptoms (GAD-7). In conclusion, the findings confirm that the online version of the PCL-5 is a valid and reliable instrument for assessing PTSD in Spanish-speaking healthcare workers, establishing it as the only DSM-5-adapted tool available for this population.

## Declarations

### Acknowledgments

The authors thank the Dirección de Investigación en Salud of the Instituto de Evaluación de Tecnologías en Salud e Investigación (IETSI) at EsSalud for their support in implementing REDCap for data collection.

### Availability of data and materials

The database can be accessed at 10.6084/m9.figshare.29143376.

### Declaration of generative AI and AI-assisted technologies in the writing process

We used DeepL to translate specific sections of the manuscript and Grammarly to improve the wording of certain sections. The final version of the manuscript was reviewed and approved by all authors.

### Ethics approval and consent to participate

The institutional research ethics committee of the National Institute of Health approved the study protocol.

### Consent for publication

Not applicable.

### Authors’ contributions

David Villarreal-Zegarra: Formal Analysis, Methodology, Supervision, Validation, Writing – Original version, Approval of the final version.

Kelly De la Cruz-Torralva: Conceptualization, Formal Analysis, Methodology, Validation, Writing – Original version, Approval of the final version.

Jaime Rosales: Recruitment coordination-Main study, Approval of the final version.

Percy Soto-Becerra: Data curation, Approval of the final version.

Ana L. Vilela-Estrada: Questionnaire curation-Main study, Approval of the final version

Cristian Diaz: Data collection-Main study, Approval of the final version

Moises Apolaya: Data collection-Main study, Approval of the final version

Rafael Leal: Data collection-Main study, Approval of the final version

Juan Barrera-Begazo: Validation, Writing – Original version, Approval of the final version.

Felipe E. García: Writing – Review & Editing, Approval of the final version.

C. Mahony Reategui-Rivera: Conceptualization, Writing – Original version, Approval of the final version.

### Acknowledgments

Not applicable.

## References

Akoglu, H. (2018). User’s guide to correlation coefficients. Turkish Journal of Emergency Medicine, 18(3), 91–93. 10.1016/j.tjem.2018.08.001

American Psychiatric Association. (2014). DSM-V: Diagnostic and Statistical Manual of Mental Disorders (5 ed.). American Psychiatric Association.

Armour, C., Műllerová, J., & Elhai, J. D. (2016). A systematic literature review of PTSD’s latent structure in the Diagnostic and Statistical Manual of Mental Disorders: DSM-IV to DSM-5. Clin Psychol Rev, 44, 60–74. 10.1016/j.cpr.2015.12.003

Armour, C., Tsai, J., Durham, T. A., Charak, R., Biehn, T. L., Elhai, J. D., & Pietrzak, R. H. (2015). Dimensional structure of DSM-5 posttraumatic stress symptoms: support for a hybrid Anhedonia and Externalizing Behaviors model. J Psychiatr Res, 61, 106–113. 10.1016/j.jpsychires.2014.10.012

Ashbaugh, A. R., Houle-Johnson, S., Herbert, C., El-Hage, W., & Brunet, A. (2016). Psychometric Validation of the English and French Versions of the Posttraumatic Stress Disorder Checklist for DSM-5 (PCL-5). PLoS One, 11(10), e0161645. 10.1371/journal.pone.0161645

Blevins, C. A., Weathers, F. W., Davis, M. T., Witte, T. K., & Domino, J. L. (2015). The Posttraumatic Stress Disorder Checklist for DSM-5 (PCL-5): Development and Initial Psychometric Evaluation. J Trauma Stress, 28(6), 489–498. 10.1002/jts.22059

Bliese, P. D., Wright, K. M., Adler, A. B., Cabrera, O., Castro, C. A., & Hoge, C. W. (2008). Validating the primary care posttraumatic stress disorder screen and the posttraumatic stress disorder checklist with soldiers returning from combat. J Consult Clin Psychol, 76(2), 272–281. 10.1037/0022-006x.76.2.272

Bovin, M. J., Marx, B. P., Weathers, F. W., Gallagher, M. W., Rodriguez, P., Schnurr, P. P., & Keane, T. M. (2016). Psychometric properties of the PTSD Checklist for Diagnostic and Statistical Manual of Mental Disorders-Fifth Edition (PCL-5) in veterans. Psychol Assess, 28(11), 1379–1391. 10.1037/pas0000254

Brewin, C. R. (2005). Systematic review of screening instruments for adults at risk of PTSD. J Trauma Stress, 18(1), 53–62. 10.1002/jts.20007

Caldas, S. V., Contractor, A. A., Koh, S., & Wang, L. (2020). Factor Structure and Multi-Group Measurement Invariance of Posttraumatic Stress Disorder Symptoms Assessed by the PCL-5. Journal of Psychopathology and Behavioral Assessment, 42(2), 364–376. 10.1007/s10862-020-09800-z

Carmassi, C., Foghi, C., Dell’Oste, V., Cordone, A., Bertelloni, C. A., Bui, E., & Dell’Osso, L. (2020). PTSD symptoms in healthcare workers facing the three coronavirus outbreaks: What can we expect after the COVID-19 pandemic. Psychiatry Res, 292, 113312. 10.1016/j.psychres.2020.113312

Carrillo-Larco, R. M., Guzman-Vilca, W. C., Leon-Velarde, F., Bernabe-Ortiz, A., Jimenez, M. M., Penny, M. E., . . . Miranda, J. J. (2022). Peru – Progress in health and sciences in 200 years of independence. The Lancet Regional Health - Americas, 7, 100148. 10.1016/j.lana.2021.100148

Cheng, P., Xu, L. Z., Zheng, W. H., Ng, R. M. K., Zhang, L., Li, L. J., & Li, W. H. (2020). Psychometric property study of the posttraumatic stress disorder checklist for DSM-5 (PCL-5) in Chinese healthcare workers during the outbreak of corona virus disease 2019. J Affect Disord, 277, 368–374. 10.1016/j.jad.2020.08.038

Di Tella, M., Romeo, A., Zara, G., Castelli, L., & Settanni, M. (2022). The Post-Traumatic Stress Disorder Checklist for DSM-5: Psychometric Properties of the Italian Version. Int J Environ Res Public Health, 19(9). 10.3390/ijerph19095282

Dosil Santamaría, M., Ozamiz-Etxebarria, N., Redondo Rodríguez, I., Jaureguizar Alboniga-Mayor, J., & Picaza Gorrotxategi, M. (2021). Impacto psicológico de la COVID-19 en una muestra de profesionales sanitarios españoles. Revista de Psiquiatría y Salud Mental, 14(2), 106–112. 10.1016/j.rpsm.2020.05.004

Durón-Figueroa, R., Cárdenas-López, G., Castro-Calvo, J., & Rosa-Gómez, A. D. L. (2019). Adaptación de la Lista Checable de Trastorno por Estrés Postraumático para DSM-5 en Población Mexicana. Acta de investigación psicológica, 9(1), 26–36.

Elhai, J. D., Biehn, T. L., Armour, C., Klopper, J. J., Frueh, B. C., & Palmieri, P. A. (2011). Evidence for a unique PTSD construct represented by PTSD’s D1-D3 symptoms. J Anxiety Disord, 25(3), 340–345. 10.1016/j.janxdis.2010.10.007

Foa, E. B., McLean, C. P., Zang, Y., Zhong, J., Powers, M. B., Kauffman, B. Y., . . . Knowles, K. (2016). Psychometric properties of the Posttraumatic Diagnostic Scale for DSM-5 (PDS-5). Psychol Assess, 28(10), 1166–1171. 10.1037/pas0000258

Franco-Jimenez, R. A., & Nuñez-Magallanes, A. (2022). [Psychometric Properties of the GAD-7, GAD-2, and GAD-Mini in Peruvian College Students]. Propósitos y Representaciones, 10(1), e1437. 10.20511/pyr2022.v10n1.1437 (Propiedades psicométricas del GAD-7, GAD-2 y GAD-Mini en universitarios peruanos)

Friedman, M. J. (2013). Finalizing PTSD in DSM-5: getting here from there and where to go next. J Trauma Stress, 26(5), 548–556. 10.1002/jts.21840

García-Campayo, J., Zamorano, E., Ruiz, M. A., Pardo, A., Pérez-Páramo, M., López-Gómez, V., . . . Rejas, J. (2010). Cultural adaptation into Spanish of the generalized anxiety disorder-7 (GAD-7) scale as a screening tool. Health Qual Life Outcomes, 8, 8. 10.1186/1477-7525-8-8

Grisanzio, K. A., Goldstein-Piekarski, A. N., Wang, M. Y., Rashed Ahmed, A. P., Samara, Z., & Williams, L. M. (2018). Transdiagnostic Symptom Clusters and Associations With Brain, Behavior, and Daily Function in Mood, Anxiety, and Trauma Disorders. JAMA Psychiatry, 75(2), 201–209. 10.1001/jamapsychiatry.2017.3951

Ito, M., Takebayashi, Y., Suzuki, Y., & Horikoshi, M. (2019). Posttraumatic stress disorder checklist for DSM-5: Psychometric properties in a Japanese population. J Affect Disord, 247, 11–19. 10.1016/j.jad.2018.12.086

Johns Hopkins University. (2022). OVID-19 World Map. Johns Hopkins Coronavirus Resource Center. Retrieved November 15, 2022 from https://coronavirus.jhu.edu/map.html

Kalkbrenner, M. T. (2023). Alpha, Omega, and H Internal Consistency Reliability Estimates: Reviewing These Options and When to Use Them. Counseling Outcome Research and Evaluation, 14(1), 77–88. 10.1080/21501378.2021.1940118

Krishnamoorthy, Y., Nagarajan, R., Saya, G. K., & Menon, V. (2020). Prevalence of psychological morbidities among general population, healthcare workers and COVID-19 patients amidst the COVID-19 pandemic: A systematic review and meta-analysis. Psychiatry Res, 293, 113382. 10.1016/j.psychres.2020.113382

Lai, J., Ma, S., Wang, Y., Cai, Z., Hu, J., Wei, N., . . . Hu, S. (2020). Factors Associated With Mental Health Outcomes Among Health Care Workers Exposed to Coronavirus Disease 2019. JAMA Network Open, 3(3), e203976–e203976. 10.1001/jamanetworkopen.2020.3976

Liu, P., Wang, L., Cao, C., Wang, R., Zhang, J., Zhang, B., . . . Elhai, J. D. (2014). The underlying dimensions of DSM-5 posttraumatic stress disorder symptoms in an epidemiological sample of Chinese earthquake survivors. J Anxiety Disord, 28(4), 345–351. 10.1016/j.janxdis.2014.03.008

Luo, M., Guo, L., Yu, M., Jiang, W., & Wang, H. (2020). The psychological and mental impact of coronavirus disease 2019 (COVID-19) on medical staff and general public - A systematic review and meta-analysis. Psychiatry Res, 291, 113190. 10.1016/j.psychres.2020.113190

Marshall, M. (2020). The hidden links between mental disorders. Nature, 581(7806), 19–21. 10.1038/d41586-020-00922-8

Mokhtari, R., Moayedi, S., & Golitaleb, M. (2020). COVID-19 pandemic and health anxiety among nurses of intensive care units. Int J Ment Health Nurs, 29(6), 1275–1277. 10.1111/inm.12800

Morrison, K., Su, S., Keck, M., & Beidel, D. C. (2021). Psychometric properties of the PCL-5 in a sample of first responders. J Anxiety Disord, 77, 102339. 10.1016/j.janxdis.2020.102339

Pfefferbaum, B., & North, C. S. (2020). Mental Health and the Covid-19 Pandemic. N Engl J Med, 383(6), 510–512. 10.1056/NEJMp2008017

Putnick, D., & Bornstein, M. (2016). Measurement invariance conventions and reporting: the state of art and future directions for psychological research. Developmental Review, 41(71-90). 10.1016/j.dr.2016.06.004

Rabow, M. W., Huang, C. S., White-Hammond, G. E., & Tucker, R. O. (2021). Witnesses and Victims Both: Healthcare Workers and Grief in the Time of COVID-19. J Pain Symptom Manage, 62(3), 647–656. 10.1016/j.jpainsymman.2021.01.139

Ruiz, M. C., Devonport, T. J., Chen-Wilson, C. J., Nicholls, W., Cagas, J. Y., Fernandez-Montalvo, J., . . . Robazza, C. (2020). A Cross-Cultural Exploratory Study of Health Behaviors and Wellbeing During COVID-19. Front Psychol, 11, 608216. 10.3389/fpsyg.2020.608216

Sahebi, A., Nejati-Zarnaqi, B., Moayedi, S., Yousefi, K., Torres, M., & Golitaleb, M. (2021). The prevalence of anxiety and depression among healthcare workers during the COVID-19 pandemic: An umbrella review of meta-analyses. Prog Neuropsychopharmacol Biol Psychiatry, 107, 110247. 10.1016/j.pnpbp.2021.110247

Schmitt, T. A., Sass, D. A., Chappelle, W., & Thompson, W. (2018). Selecting the "Best" Factor Structure and Moving Measurement Validation Forward: An Illustration. J Pers Assess, 100(4), 345–362. 10.1080/00223891.2018.1449116

Seligowski, A. V., & Orcutt, H. K. (2016). Support for the 7-factor hybrid model of PTSD in a community sample. Psychol Trauma, 8(2), 218–221. 10.1037/tra0000104

Shaukat, N., Ali, D. M., & Razzak, J. (2020). Physical and mental health impacts of COVID-19 on healthcare workers: a scoping review. Int J Emerg Med, 13(1), 40. 10.1186/s12245-020-00299-5

Simms, L. J., Watson, D., & Doebbeling, B. N. (2002). Confirmatory factor analyses of posttraumatic stress symptoms in deployed and nondeployed veterans of the Gulf War. J Abnorm Psychol, 111(4), 637–647. 10.1037//0021-843x.111.4.637

Singh, R., & Subedi, M. (2020). COVID-19 and stigma: Social discrimination towards frontline healthcare providers and COVID-19 recovered patients in Nepal. Asian J Psychiatr, 53, 102222. 10.1016/j.ajp.2020.102222

Spitzer, R. L., Kroenke, K., & Williams, J. B. (1999). Validation and utility of a self-report version of PRIME-MD: the PHQ primary care study. Primary Care Evaluation of Mental Disorders. Patient Health Questionnaire. Jama, 282(18), 1737–1744.

Spitzer, R. L., Kroenke, K., Williams, J. W., & Löwe, B. (2006). A brief measure for assessing generalized anxiety disorder: The GAD-7. Archives of Internal Medicine, 166(10), 1092–1097. 10.1001/archinte.166.10.1092

Suh, Y. (2015). The Performance of Maximum Likelihood and Weighted Least Square Mean and Variance Adjusted Estimators in Testing Differential Item Functioning With Nonnormal Trait Distributions. Structural Equation Modeling: A Multidisciplinary Journal, 22(4), 568–580. 10.1080/10705511.2014.937669

Sun, J. (2005). Assessing Goodness of Fit in Confirmatory Factor Analysis. Measurement and Evaluation in Counseling and Development, 37(4), 240–256. 10.1080/07481756.2005.11909764

Tsai, J., Harpaz-Rotem, I., Armour, C., Southwick, S. M., Krystal, J. H., & Pietrzak, R. H. (2015). Dimensional structure of DSM-5 posttraumatic stress disorder symptoms: results from the National Health and Resilience in Veterans Study. J Clin Psychiatry, 76(5), 546–553. 10.4088/JCP.14m09091

Villarreal-Zegarra, D., Barrera-Begazo, J., Otazú-Alfaro, S., Mayo-Puchoc, N., Bazo-Alvarez, J. C., & Huarcaya-Victoria, J. (2023). Sensitivity and specificity of the Patient Health Questionnaire (PHQ-9, PHQ-8, PHQ-2) and General Anxiety Disorder scale (GAD-7, GAD-2) for depression and anxiety diagnosis: a cross-sectional study in a Peruvian hospital population. BMJ Open, 13(9), e076193. 10.1136/bmjopen-2023-076193

Villarreal-Zegarra, D., Copez-Lonzoy, A., Bernabe-Ortiz, A., Melendez-Torres, G. J., & Bazo-Alvarez, J. C. (2019). Valid group comparisons can be made with the Patient Health Questionnaire (PHQ-9): A measurement invariance study across groups by demographic characteristics. PLoS One, 14(9), 1–15. 10.1371/journal.pone.0221717

Wolf, E. J., Mitchell, K. S., Sadeh, N., Hein, C., Fuhrman, I., Pietrzak, R. H., & Miller, M. W. (2017). The Dissociative Subtype of PTSD Scale: Initial Evaluation in a National Sample of Trauma-Exposed Veterans. Assessment, 24(4), 503–516. 10.1177/1073191115615212

Wu, H., & Estabrook, R. (2016). Identification of Confirmatory Factor Analysis Models of Different Levels of Invariance for Ordered Categorical Outcomes. Psychometrika, 81(4), 1014–1045. 10.1007/s11336-016-9506-0

Wu, K. K., Lee, D., Sze, A. M., Ng, V. N., Cho, V. W., Cheng, J. P., . . . Tsang, O. T. (2022). Posttraumatic Stress, Anxiety, and Depression in COVID-19 Survivors. East Asian Arch Psychiatry, 32(1), 5-10. 10.12809/eaap2176

Xiong, J., Lipsitz, O., Nasri, F., Lui, L. M. W., Gill, H., Phan, L., . . . McIntyre, R. S. (2020). Impact of COVID-19 pandemic on mental health in the general population: A systematic review. J Affect Disord, 277, 55–64. 10.1016/j.jad.2020.08.001

Zhou, T., Huang, S., Cheng, J., & Xiao, Y. (2020). The Distance Teaching Practice of Combined Mode of Massive Open Online Course Micro-Video for Interns in Emergency Department During the COVID-19 Epidemic Period. Telemed J E Health, 26(5), 584–588. 10.1089/tmj.2020.0079

